# Measurement of 24-hour Continuous Human CH_4_ Release in a Whole Room Indirect Calorimeter

**DOI:** 10.1101/2022.11.04.22281777

**Authors:** E.A. Carnero, C.P. Bock, Y. Liu, K.D. Corbin, E. Wohlers-Kariesch, K. Ruud, J. Moon, M. Andrew, R. Krajmalnik-Brown, A. Muraviev, K.L. Vodopyanov, S.R. Smith

## Abstract

We describe the technology and validation of a new whole room indirect calorimeter (WRIC) methodology to quantify methane (CH_4_) released from the human body over 24h concurrently with the assessment of energy expenditure and substrate utilization. The new system extends the assessment of energy metabolism by adding CH_4_, a downstream product of microbiome fermentation that could contribute to energy balance.

**Methods:** Our new system consists of an established whole room indirect calorimeter WRIC combined with the addition of off-axis integrated-cavity output spectroscopy (OA-ICOS) to measure CH_4_ concentrations ([CH_4_]). The volume of CH_4_ released (VCH_4_) was calculated after measuring air flow rates. Development and validation included environmental experiments to measure the stability of the atmospheric [CH_4_], infusing CH_4_ into the WRIC and cross-validation studies comparing [CH_4_] quantified by OA-ICOS and mid-infrared dual-comb spectroscopy (MIR DCS). Reliability of the whole system is reported between years, weeks, days, and validated CH_4_ infusions. The cross-validation and reliability of VCH_4_ released from the human body was determined in 19 participants on consecutive days. In addition, we describe a postprocessing analytical method to differentiate CH_4_ released from breath versus intestine by matching times of stool production and contemporaneous VCH_4_ release.

**Results:** Our infusion data indicated that the system measured 24h [CH_4_] and VCH_4_ with high sensitivity, reliability and validity. Cross-validation studies showed good agreement between OA-ICOS and MIR DCS technologies (r= 0.979, P<0.0001). Initial human data revealed 24h VCH_4_ was highly variable between subjects and within / between days; this highlights the importance of a 24-h continuous assessment to have a complete picture of VCH4 release. Finally, our method to quantify VCH_4_ released by breath or colon suggested that over 50% of the CH_4_ was eliminated through the breath.

**Conclusions:** The method allows, for the first time, measurement of 24h VCH_4_ (in kcal) and therefore the measurement of the proportion of human energy intake fermented to CH_4_ by the gut microbiome and released via breath or directly from the intestine. Our method is accurate, valid, and will provide meaningful data to understand not only interindividual variation, but also allows us to track the effects of dietary, probiotic, bacterial and fecal microbiota transplantation on VCH_4_.

## Introduction

Quantification of human metabolism is fundamental to metabolic disease research and Whole Room Indirect Calorimeters (WRIC) are important tools in the study of metabolism. To date, the field of indirect calorimetry is focused largely on the measurement of Energy Expenditure (EE), substrate oxidation, spontaneous physical activity and more recently metabolic flexibility (1, 2).

Energy balance research typically consists of 24h or longer studies in metabolic units often including WRIC to measure Energy Expenditure (EE). Energy consumed is observed and duplicate meals are quantified in a food lab representing Energy Intake (EI). Energy balance is often calculated as simply EI – EE with an assumed fixed proportion of the food digested and available to the human host. Urinary energy loss (mostly urea nitrogen and glucose in diabetes) are also important to the energy balance equation (3). Components of energy balance that are less studied are metabolizable energy (4) and the energy lost as methane (4, 5). Important to this investigation, methane production (VCH_4_) has not been incorporated in energy balance research (5).

The microbiome ferments colonic carbohydrates to provide substrate to at least three species of archaea in the gut microbiome to produce methane gas (6). Importantly, methane production varies from person to person. The existing literature, based on single breath analysis, reveals that there are two distinct populations of methane ‘producers’ (6, 7) and the proportion of ‘producers’ varies widely across populations (6, 8). Continuous 24-hour monitoring of methane production has rarely been done in humans (7) and never under ambulatory environmental conditions (walking, sleeping). Single breath tests may capture only 20% of daily CH_4_ produced by the human gut microbiome (8). Some humans do not release CH_4_ through the breath, although they have significant colonic production (9).

CH_4_ is produced only by gut microbes and its presence indicates microbial activity (9-11). CH_4_ trapped in the feces and flatulencies are the main routes to release CH_4_. Fecal CH_4_ is released via the anal sphincter after defecation (10). The pathway to release CH_4_ through the breath is more complicated and involves active transport through the colon enterocytes, passage to and transport in the portal, systemic and lung circulation, and finally transport through the lung interstitium and alveoli.

While CH_4_ is considered an inert end-product of intestinal microbial processes (11), animal studies have suggested CH_4_ functions as a neuromuscular transmitter to delay intestinal transit (12). Independent of CH_4_ function, it has been suggested that CH_4_ may exist in steady state and fluctuate in response to physiological events like digestion after meals or exercise (13). Also, sampling CH_4_ over time has been proposed to be a superior diagnostic method for CH_4_ producers than single breath tests (14).

Given the potential role of CH4 in human energy balance, physiology and disease diagnosis, more reliable and comprehensive methods to quantify CH4 production/release from human microbiome are necessary. Therefore, the main goals of this paper were to describe the setup and validation of a system to measure continuous human gut microbiome CH4 production integrated in a WRIC for human studies. In addition, we aimed to create a postprocessing gas analysis to differentiate between colonic and breath CH4 release.

## Method

### Participants

Participants included in this manuscript were enrolled as part of a previous clinical trial (NCT02939703) approved by the AdventHealth Institutional Review Board and conducted at the AdventHealth Translational Research Institute. The complete methods from that study have been described previously (5). All potential participants provided written informed consent. Briefly, the study recruited healthy, weight stable males and females, age 18-45 years, BMI ≤ 30 kg/m^2^. The topline results of that trial are forthcoming. Therefore, here, we used data from participants who only completed a portion of the study. All individual days of each participant inside the WRIC were included validation studies and post-processing analysis. For the initial cross-validation studies, the participants were specifically recruited for the purpose of the experiments (IRB: #321421). Additional information about these two protocols is provided in the specific section below within the context of our analytical approach.

### Design

This manuscript describes a series of technology procedures, gas exchange experiments and, validation and reliability human studies with the goal of installing a CH_4_ analysis system integrated into a WRIC. Firstly, we conducted several experiments to analyze the environmental fluctuations of CH_4_ to test the stability of inflow methane coming from our medical air system which includes a buffer tank of 1,514.165 L at 100 lb/in^2^. Then, we performed studies of validity and reliability of the integrated system to measure CH_4_ release inside the WRIC with gas infusions. Afterwards, we cross-validated CH_4_ concentrations from our system with a reference method and finally, we used a sample of 19 participants to collect repeated measures of 24-hour continuous CH_4_ volume to assess the reliability of human CH_4_ release. Environmental experiments, internal validation and reliability, human cross-validation and human reliability studies are described in detail in the next sections.

### Environmental Experiments

The WRIC system includes sequential components to measure and control gas flow: the inflow, the chamber and the analyzer. To introduce a new analyzer into the calorimetry system at the end of that sequence, we needed to measure methane at all exchange and accumulation points. Methane has a reported environmental background of 1.8 ppm (11), but the hourly and daily fluctuations are dictated by many factors including wind and traffic patterns, and establishments nearby that may be large CH_4_ producers (petrochemical dispensing stations, sewage treatments plants, landfills, swamps, etc.).

To analyze the stability of our inflow gases, we first measured CH_4_ from the medical air system (Figure 1 a). We then measured below the roof line of the research building on the three sides nearest the medical air equipment look for the most stable point to measure CH_4_ (Figure 1 b). Our medical air system is housed in a rooftop structure on the north side of the building. The three closest sides where we could feasibly plumb a new inlet were east, with a petrochemical station as an immediate neighbor, north, which is over the building back lot with a diesel power generator, mass N_2_ storage as well as sewer vents and west, which has no direct neighbor, but an interstate highway ∼140 m away. Daily methane patterns are shown in Figure 1 a-b. The inlet for the medical air system was originally plumbed to above the roof of the building. Based on this data we used the West piping as the air inlet.

**Figure 1.**
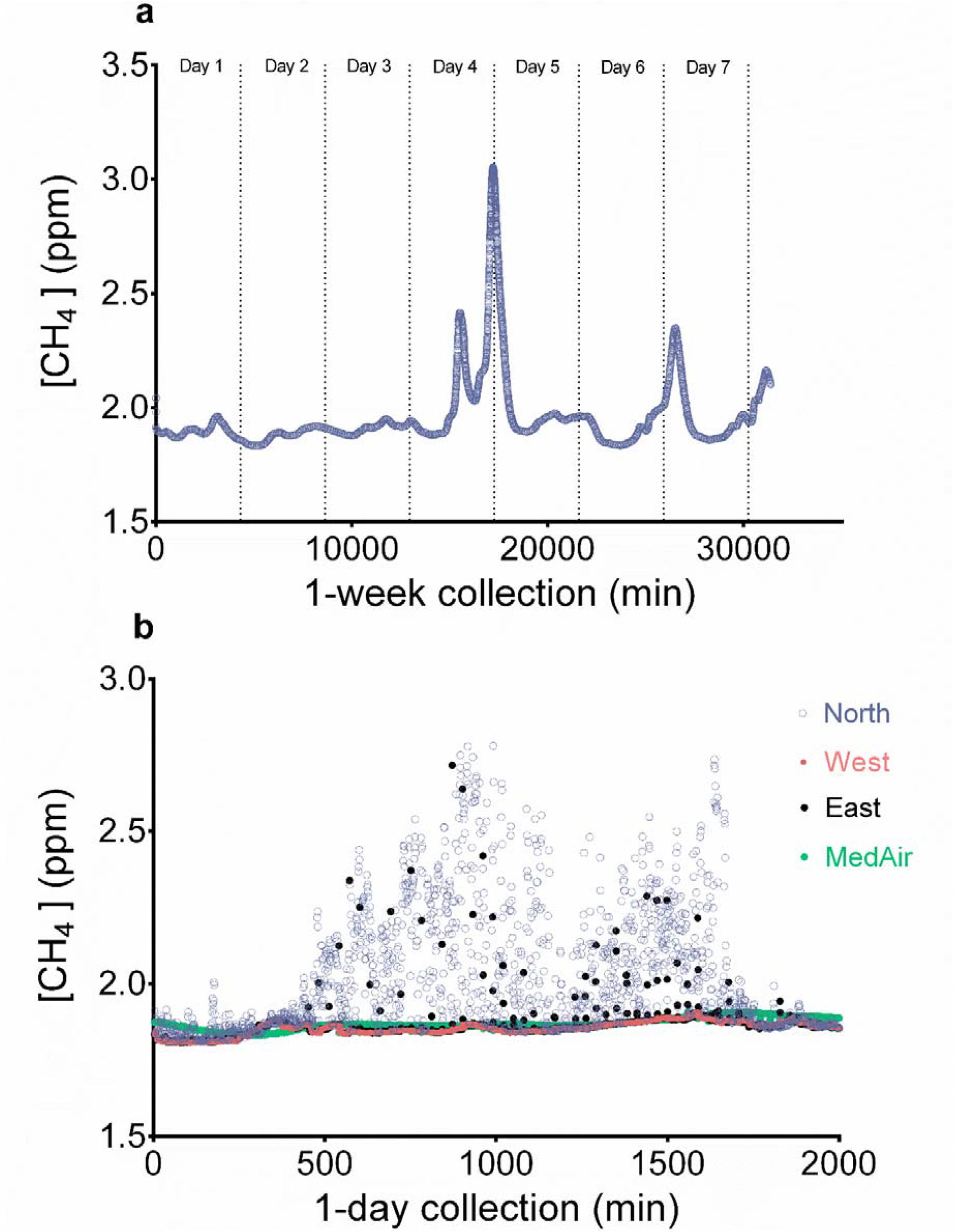
Variability of environmental methane concentration ([CH_4_]) as function of the air inflow pipeline location. (a) [CH_4_] assessment over one week, CH_4_ was measured with an off-axis integrated-cavity output spectroscopy (OA-ICOS). (b) [CH_4_] measured simultaneously during 1-day on 3 different environmental locations and from the reference air tank.

### Validation Studies

#### Internal validation and reliability (instrument validation)

After we established the most stable inlet for methane, the next step was to validate the methane analyzer (off-axis integrated-cavity output spectroscopy (OA-ICOS)) using a National Institute of Standards and Technology (NIST) traceable tank of 40 ppm CH_4_ methane with the balance of the constituents consisting of environmental air. Prior to validating the analyzer within the system, we first infused gas (dried environmental air and 40 ppm CH_4_) directly to the analyzer using a mass flow controller (MFC). The measured concentration remained at 40 ppm with multiple flow rates. Next, infusion studies were performed to validate the WRIC system as a tool to measure methane production against established methods. Standard quality control procedures for the WRIC, involve blender-controlled infusions of nitrogen (N_2_) and Carbon Dioxide (CO_2_) to simulate human metabolism (1). These infusions are performed into the chamber as well as using a short plumbing system that bypasses the chamber, this “short circuit” check allows for quick troubleshooting by removing the volume factor both in terms of infusion time and within the calculations. These same methods were applied using the CH_4_ mixed gas (40 ppm CH_4_) by first infusing into the short circuit and then to the chamber. We then measured the output gases with the methane analyzer.

The short circuit infusion was completed over 40 min with three different flow rates (1 L/min, 2 L/min, and 3 L/min) with a constant inflow rate of 60 L/min. The second set WRIC infusions were performed over 18 hours with the first 10 hours at 1 L/min and the last 8 hours at 0.75 L/min. There were also two 15-minute jumps to 3 L/min at the 4-hour mark and the 8.25-hour mark (these jumps were intended to simulate fast changes in CH_4_ production during flatulence). Expected and measured VCH_4_ was calculated, and overall recovery used to determine error (Figures 2 a-f). In the last set of internal validation procedures, we performed series of short step infusions (10 minutes per step at 0.1% [CH_4_]) simulating from the lowest (0.0083 mL/min) to high (0.1388 mL/min) expected volumes of human CH_4_ release (the first series in progressive and the second in random order); this procedure permitted us to validate small changes in CH_4_ release in a short period of time (Supplemental Figure 1).

**Figure 2.**
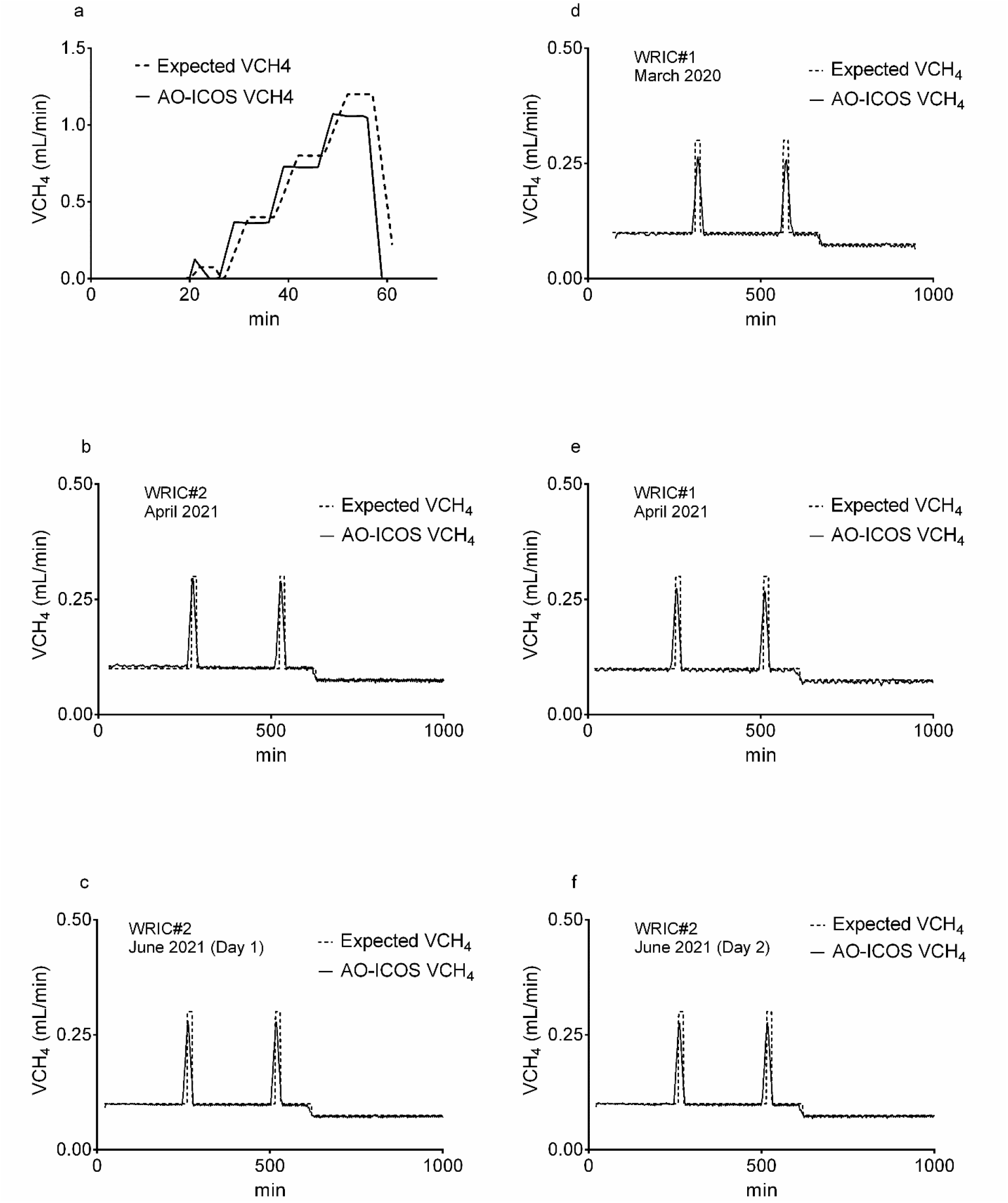
**a-f**. Methane volume (VCH_4_) measured during 1,000-minute infusion studies in whole room indirect calorimeter (WRIC). Dashed lines represent expected CH_4_ volumes infused with a blender and measured by flow mass controllers (MFC); solid lines are CH_4_ volumes measured by off-axis integrated-cavity output spectroscopy (OA-ICOS). Figure 2a shows a short circuit dynamic infusion. Figures 2 b-f are 2-step dynamic infusion in two different WRIC (#2 and #1) at the same time point (b and e); in the same WRIC (#1) one year apart (e and d); in the same WRIC (#2) in 2 consecutive days (c (day1) and f (day2)).

#### Cross-validation

To cross-validate our measurements of methane concentration with the OA-ICOS analyzer, we collaborated with the MirCombs Research Group at the College of Optics and Photonics at the University of Central Florida (CREOL, UCF) to measure methane concentration with another technique of broadband mid-infrared frequency comb laser spectroscopy (see section: CH_4_ measurement by middle infra-red dual-comb spectroscopy (MIR DCS)). We collected 47 single time point gas samples from the outflow of the WRIC in gas sampling bags. The delay time between sample collection and the OA-ICOS measurements was negligible; therefore, methane concentrations were measured with the two different techniques at the same time to within a minute.

#### Human Reliability Studies

A test-retest approach was used by repeating 24-hour measurements in the WRIC on consecutive (between day reliability) and non-consecutive (between-week reliability) days. This study design permitted us to assess our ability to reproduce a test without the instrument introducing potentially misleading errors (see validation studies above). All assessments were preceded by a 3-day stay at the Translational Research Institute AdventHealth Clinical Research Unit (CRU). Participants were admitted at 18:00 and consumed all 3-day meals provided by the TRI metabolic kitchen at the CRU. After a 12-hour overnight fasting period (third night at TRI) participants were transferred to the WRIC to start the metabolic assessments. All WRIC assessments were conducted in energy balance and followed the same schedule of activities and diet between-day and weeks (Supplemental Figure 2). Seven days were utilized to calculate the overall CH_4_ release reliability; six of the 7 days were consecutive and were used to calculate the between-day reliability and the between-week reliability was calculated comparing the mean of 6 consecutive days with the 7^th^ day (two weeks apart).

### Whole Room Indirect Calorimetry

#### Description of the Basics the Whole Room Indirect Calorimeter (WRIC)

The WRIC setup provides precise measures of VO_2_ and VCO_2_ by using a controlled testing environment. Two WRIC chambers with volumes of 31,000 L were used to perform 24-hour energy expenditure (24-hour EE) studies with participants staying overnight for 23-hours. We then extrapolate these values to 24-hour (value of 23-hour + 1 hours of mean 23-hour value). Simulating free living through scheduled activities allows for reproducible energy expenditure measurements (5). WRIC use mass flow controllers (MFC) for inflow and outflow that can be analyzed by O_2_ and CO_2_ gas analyzers. Our WRIC is a push-pull system with a reference air system serving as the inflow air (push) and a pull MFC to control gas concentrations for operation at lower pressure and more stable data.

#### Reference Air System

The use of a reference air system from a large buffer tank minimizes changes in gas concentrations within the WRIC as atmospheric air composition is known to change throughout the day. Our reference air system uses two tanks to reduce instrument noise and produce a large buffer tank of dry air as the draw point for the inflow air. The system is located in a machine room penthouse on the roof of our facility with the inlet pipe on top of the structure to the west. Environmental concentrations of oxygen and carbon dioxide are larger portions of air relative to volatile organic compounds (VOC) and are therefore much more consistent. Methane, with a current global concentration of 1850 ppb (11), can be greatly impacted if there is a local source temporarily present near the building. To this end, we performed an analysis of the four sides of the building (describe in the previous section) to determine which direction had the most consistent data and lower variability.

#### VO_2_, VCO_2_ and CH_4_ Gas Analyzers

Inflow and outflow (chamber sample) O_2_ and CO_2_ were continuously measured by dual channel Ultramat/Oxymat 6 Siemens gas analyzers. The oxygen analyzer is a paramagnetic sensor with a 5ppm resolution, and the carbon dioxide analyzer is an infrared sensor with a resolution of 1ppm. The analyzers use a reference gas of ∼21% O_2_ with the balance N_2_ at a very low flow rate through the analyzers (10 mL/min). A software calibration is performed within CalRQ software (MEI Research, MN, USA) when the delta between the zero-calibration gas and the span calibration gas is not within ±3% of the expected delta. A hardware calibration is performed anytime a calibration check exceeds that mark and at every reference gas tank change. A null correction is also performed by running medical air into both the inflow analyzer and the WRIC analyzer prior to each study and then aligning O_2_ and CO_2_ in post processing.

#### Push-Pull, Proportional–Integral–Derivative Controller and Push Math

Precise measurement of VOCs requires control over the inflow air quantity (flow rate) and quality (stability of gas concentrations). The push configuration paired with proportional–integral–derivative controller (PID controller) is the best choice to maximize these three goals. Measuring gas concentrations on the inflow (push) side while maintaining positive pressure in the chamber, allows for control over all air that enters the chamber. Flow rate will be dictated by carbon dioxide levels in the chamber allowing the rate to remain as low as possible and increasing the ability to capture a change in gas concentrations at the parts per billion level.

CalRQ *is* the software that was built for these calorimeters by the engineers that designed and built the WRIC (TRI-AH-CRU). The software CalRQ runs the system and records all data streams as well as performing live processing for the graphs. The software can be set up to record in averaging intervals of 15, 30 or 60 sec depending on the resolution necessary. Normal operation including all the assessments included herein were performed using the 60 sec intervals. The software records O_2_, CO_2_, humidity for both inflow and outflow air along with temperature, humidity and pressure for both the chamber and the instrumentation room. The data set also includes inflow rate and activity. The software calculates VO_2_ and VCO_2_ with the following equations:

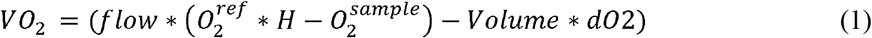

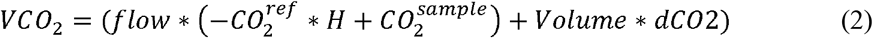

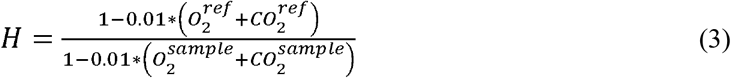

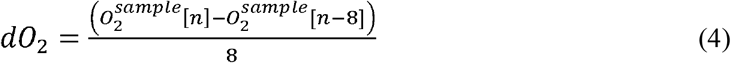

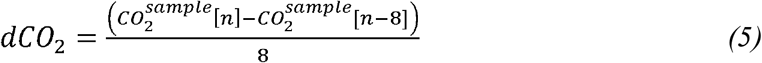

*Where ref is the inflow air from the med air unit and sample is the outflow air sampled from the chamber. H is Haldane correction. n, represents the minute of sample collection (there are 1380 minutes collected per day). Number 8, it is the length of the derivate in minutes. The derivative terms (dO*_*2*_ *and dCO*_*2*_*) are changed from the backward derivative seen above to a centered derivative below in post processing*.

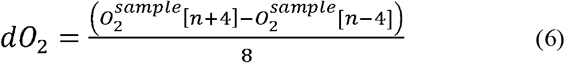

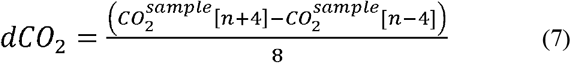

After post processing, the resulting VO_2_ and VCO_2_ data can be used to calculate energy expenditure (EE) and respiratory exchange ratio (RER = VCO_2_/VO_2_). EE (kcal/24-hour) was calculated from VO_2_, VCO_2_, VCH_4_ and Urinary Nitrogen (UN) if collected using an appropriate equation (3). VCH_4_ measurements were obtained by an additional analyzer (see next section) and were aligned in time with VO_2_ and VCO_2_.

#### Methane Analysis and Data Processing

There are space limitations when adding a secondary analyzer into the WRIC system, in this instance methane; a need to minimize cost and avoid an impact on the primary purpose of the system. To measure methane in our system, we selected a Los Gatos Research (LGR) Greenhouse Gas Analyzer, which measures, in a single device, CH_4_, CO_2_ and H_2_O simultaneously. The analyzer is based on OA-ICOS technology, a fourth-generation cavity enhanced laser absorption technique, which allows for ultra-stable and fast analysis. The principal performance specifications of this analyzer are: Precision (100 seconds): 0.3 ppb, maximum drift: 5 ppb, accuracy: <1%, measurement range (100 seconds): 0.01-100 ppm, operational range: 0-1000 ppm.

Paired with an LGR Multi Inlet Unit (MIU), this analyzer allowed us to measure all WRIC (a total of 4 rooms) as well as inflow air. OA-ICOS uses an infrared pathlength of many kilometers to be able to measure small volatiles at low (part per billion) concentrations in seconds; the instrument provides CH_4_ and CO_2_ on a dry (and wet) mole fraction without any need to dry samples. Each inlet to the MIU is plumbed from the instrumentation rack after drying and prior to the O_2_ and CO_2_ analyzers to create parallel analyzer paths. The MIU allows the lines to remain under pressure while switching through the inlets using 4 minutes for the chambers and 2 minutes for the inflow air. This setup met our needs of space, cost and zero interference with the primary endpoint (usually, VO_2_ and VCO_2_).

The analyzer saves raw files locally as well as feeding live data into the CalRQ software that operates the WRIC. The live data allows real time monitoring and troubleshooting for gas measurements from the chamber, but the most accurate analysis requires post processing. While VO_2_ and VCO_2_ require a null correction and centering the derivative in post processing, CH_4_ data must go through several post processing steps before a derivative term can be used to calculate VCH_4_. The raw methane data (ppm) is stored in 5-hour files of second-by-second data with the inlet channel specified within the data set. The data is sorted by channel, averaged to minute-by-minute data and then interpolation is used to fill the gaps. After all blocks of data are processed for a day, the data are combined into a single file and converted into percent inflow and outflow values. These values are then paired with the calorimetry data and VCH_4_ is calculated using a 16-min derivative (equation 8 and electronic supplementary information).

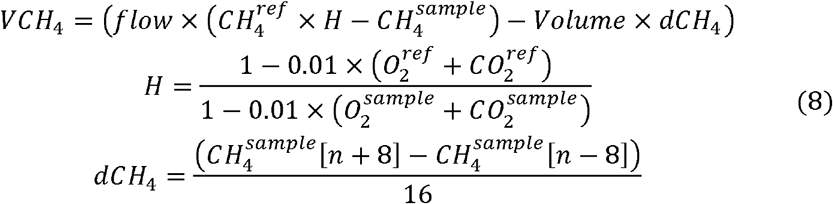

*Where ref is the inflow air from the med air unit and sample is the outflow air sampled from the chamber. The derivative term (dCH*_*4*_*) uses a centered derivative, and it is applied in post processing. n, represents the minute of sample collection (there are 1380 minutes collected per day). Number 16, it is the length of the derivate in minutes*.

A standard calibration curve for the whole system (WRIC + OA-ICOS) was created from low to high concentrations to confirm the validity of the system to measure low (< 50 ml/day) and high (>50 ml/day) CH_4_ producers. Short steady-state CH_4_ infusions were performed every 20 minutes starting at concentrations equivaling to 25 ml/day production and following with 50 ml/day, 100 ml/day, 250 ml/day, 500 ml/day, 1000 ml/day and 1500 ml/day productions.

### CH_4_ measurement by middle infra-red dual-comb spectroscopy (MIR DCS)

For cross validation of CH_4_ concentration measurements, we used gas samples from the outflow of the WRIC that were collected in sampling bags and analyzed them at CREOL, UCF using mid-infrared dual-comb spectroscopy (MIR DCS) – currently the most advanced spectroscopic technique for precise and massively parallel measurements of trace gas concentrations. The MIR DCS system shown in Supplement Figure 3(a) and described in detail in (15) used a pair of broadband (instantaneous spectral coverage 3.2–5.4 μm) frequency combs based on subharmonic generation in optical parametric oscillators (OPOs) pumped by two phase-locked Tm-fiber lasers. The DCS method combines high spectral resolution, high detection sensitivity and the capability of simultaneous detection of multiple species (including isotopologues), thanks to its broadband spectral coverage. The instrument features the absolute precision of the optical frequency scale through referencing it to an atomic clock and high accuracy of the absolute gas concentration measurement through comparing the intensities of multiple spectral lines with those from molecular spectral libraries, such as HITRAN. Gas from the sampling bags was routed into a compact multipass optical cell. A vacuum pump, pressure-control valve, and an external controller were used to maintain the desired working pressure (10–30 mbar) in the cell. The reduced pressure was used to avoid collisional broadening of spectral lines and achieve good visibility of spectral features. After each set of measurements, the gas was exhausted from the multipass cell via a vacuum pump. Supplement Figure 3(b) shows the MIR DCS spectrum of methane near 3.26-μm wavelength (3067 cm^-1^) along with a simulated spectrum from the HITRAN database (inverted). The absolute concentration of CH_4_ was determined via matching the heights of the measured and simulated spectral peaks. The MIR DCS system allowed part-per-billion level of detection sensitivity for methane and about 1% fractional accuracy of its absolute concentration measurement. This high accuracy was achieved because of involving multiple absorption peaks in the fitting procedure.

### Post-Processing Data Analysis

#### CH4 released by Breath and Intestine

After measuring and processing the 24-hour [CH_4_] output/raw data, we obtained a minute by minute 24-hour VCH_4_ profile. From stool collection times (see next section), we were able to detect peaks of CH_4_ associated with colonic release. In addition, we included in the intestinal production-colonic released (from here on out, colonic released VCH_4_) calculation every peak/area in a range above a localized baseline steady state (VCH_4FILTERED_). We determined a baseline steady state during sleep (0:00-6:00 hours) after excluding peaks above the baseline related to movement and considering the OA-ICOS variability. We assumed as intestinal VCH_4_ release any value above the mean sleeping value times the coefficient of variation (CV) of all individual sleeping minutes filter to movement plus technical error of OA-ICOS device variation (calculated from CV of static infusions) times 24-hour VCH_4_. The formula for calculating colonic released VCH_4_ is therefore:

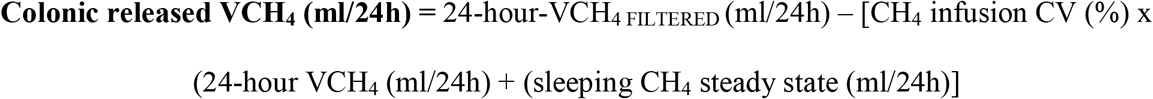

### Stool Collections

Stool collection methodologies are described elsewhere within the context of the parent study NCT02939703 and referenced elsewhere (5). Briefly, fecal samples were collected during 24-hour WRIC stays. The toilet inside the chamber was setup with specific containers for fecal collection. After depositing the sample, participants capped the container and alerted WRIC technician so that the sample could be transferred to the lab. Samples were processed in an anaerobic chamber within 1 hour of being produced for multiple downstream applications. Long term-storage of samples was at −80°C until further analysis. A stool collection time log was used to document all samples collected during the stays in the building.

### Statistical Analysis

Sample characteristics of the participants are given as mean and standard deviations. Mean [CH_4_] from environmental experiments from the 4 distinct locations were reported as 24-h area under the curve (AUC) and variabilities were compared across locations.

The accuracy of infused VCH_4_ versus measured VCH_4_ (OA-ICOS) values from internal validation infusion studies was obtained from bias correction factor (Cb) by carrying out concordance correlation coefficient analyses. Precision was described with Pearson’s coefficient correlation (r). Reproducibility between both methods was studied using the concordance coefficient correlation (ρ_c_). Simple linear regression between pairs of measurements was used to confirm significant association between VCH_4_ between measurements, as well individual slopes and intercepts constants of each regression were compared with 45 degrees line (concordance coefficient correlation, ρ_c_) and zero intercept respectively (16). In addition, an agreement analysis was conducted to confirm systematic and proportional bias by using Bland and Altman plots (17) and Kendall’s Tau correlation coefficients. The same procedure was used with the cross-sectional validation of human data to compare [CH_4_] between OA-ICOS and mid-IR DCS data.

Reliability between days and weeks for 24-h and sleep VCH_4_ was calculated as the mean differences, coefficient of variation (CV = (√((∑(test1-test2)^2^)/2N) and relative CV (%CV, (CV/mean)), respectively. All statistical analyses were carried out using (JMP 13.2.1 [SAS Institute, USA] and Prism 6.07 [GraphPad Software, USA]).

## Results

### Environmental Experiments

Measurements of environmental CH_4_ were collected at the roof of our building over a one-week period with significant fluctuations across days (Figure 1a). Afterwards, environmental [CH_4_] was measured simultaneously in 3 different locations of our building (West, East and South). Profiles of the 24-h [CH_4_] differed significantly across locations (Figure 1b). West location was the one with the lowest concentration measured and variability (West, 1.19%; East, 2.74%; North, 9.86%) and AUC (West, 3770; East, 3784; North, 4069) in [CH_4_] along the 24-hour cycle and it was selected as the final location (orientation at 254-degree W at 27 meters of altitude) for environmental gas collector to feed our medical air tank.

### Internal Validation Data

Initially, a series of short-circuit dynamic infusions were carried out to explore the capabilities of the WRIC and OA-ICOS to detect the CH_4_ signal. After these initial infusions, we observed VCH_4_ peaks in agreement with the VCH_4_ injected with the blender inside the WRIC (Figure 2a). We also performed five, 2-step CH_4_ static infusions of 5 and 10 liters (Figure 2b-f). AUCs of expected (blender) and measured (AO-ICOS) VCH_4_ values overlapped for all infusions (Figure 2b, 99.0 vs. 99.7 mL/min; Figure 3c, 96.6 vs. 94.4 mL/min; Figure 2d, 86.7 vs. 83.4 mL/min; Figure 2e, 96.6 vs 93.3 mL/min; Figure 3f, 96.6 vs. 94.7 mL/min). In addition, the comparison between VCH_4_ measures from infusions performed one year apart showed a CV of 11.6% (0.0449 mL/min) and high precision (r = 0.925, *p<0.001*). The infusions carried out in different WRIC during the same period (the same week) showed a smaller CV of 6.4% (0.0249 mL/min) and higher precision (r = 0.983, *p<0.001*) than comparison between years. The lowest CV and the highest precision were observed in the infusions performed in the same WRIC in consecutive days (Figures 2c and 2e), 1.8% and r=0.997 (*p<0.001*), respectively. Regarding the within 2-minute variability for the same infusion, the CV was 3.1% (0.0121 mL/min). This constitutes the minimum variability expected when measuring continuous VCH_4_ and it is due to technical error of AO-ICOS in the WRIC setup. Finally, our standard calibration curve built with short step infusions showed linearity (progressive infusion) and, steady and fast response (progressive and random infusions) values between infused concentrations and measured by the WRIC-AC-ICOS system (Supplementary Figure 1).

**Figure 3.**
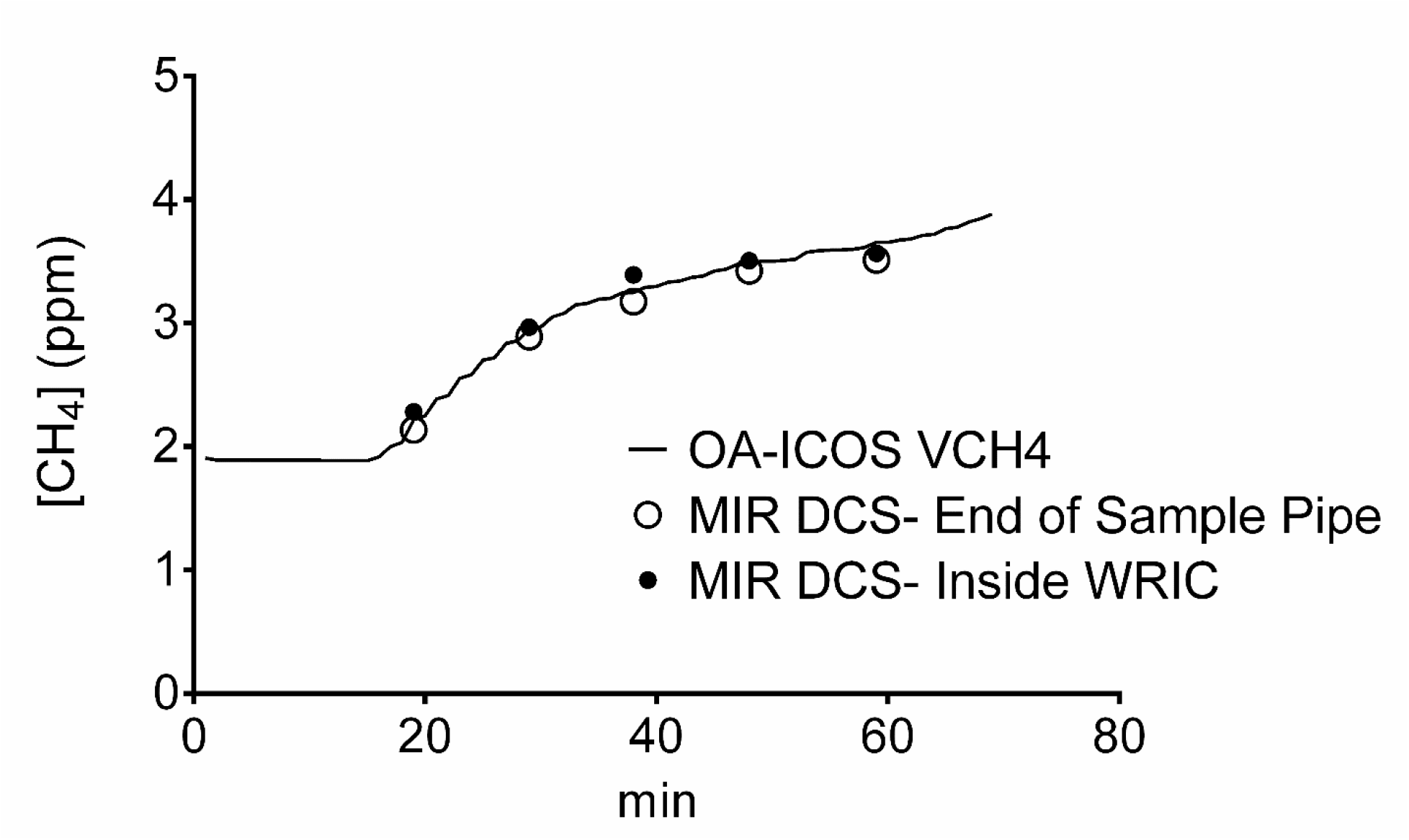
Comparison of human-microbiome methane concentration ([CH_4_]) production by different measurement methods. Solid black line represents [CH_4_] quantified directly from the air outflow of a room indirect calorimeter (WRIC) by off-axis integrated-cavity output spectroscopy (OA-ICOS) CH_4_ analyzer. The open and closed circles represent measurements done with broadband mid-infrared laser spectroscopy (MIR DCS) at the end of the line prior to entering the multi-inlet unit (MIU) and inside the WRIC, respectively.

### Cross Sectional Validation

An initial experiment with one participant was performed to confirm there were no leaks along the sample pipeline. After the first 30 minutes inside the WRIC, two sample bags were collected every 30 minutes over a period of two hours, one bag was collected inside the WRIC and another at the end of the line before entering in MIU (Figure 3). The bagged gas samples were analyzed by MIR DCS and compared to the OA-ICOS. No significant differences in [CH_4_] were observed between sample collection sites or analyzers. The [CH_4_] values in bagged samples or measured continuously by the OA-ICOS followed a similar pattern (Figure 3).

The second cross-sectional validation study was performed with random samples of 19 healthy subjects who participated in energy expenditure assessments (see Table 1 for sample characteristics). The forty-seven single time point gas samples collected from the outflow of the WRIC collected in gas sampling bags revealed good accuracy and high precision between MIR DCS and OA-ICOS (*Cb = 0.857* and r= 0.979, *p<0.001*). The concordance coefficient correlation was moderate (ρ_c_= 0.839) and the slope of the regression line between both methods was significantly different from the identity line (*p<0.0001*, Figure 4). The concordance study was completed by performing an agreement analysis with a Bland and Altman plot, which showed difference between MIR DCS and OA-ICOS of –0.400±591 ppm. Also, there was a significant correlation between the mean and the difference between methods, which indicated systematic and proportional bias, respectively (Figure 5). However, the visual inspection of the data indicates that the most important differences are found at high concentrations above 4 ppm. Altogether, these results indicate that OA-ICOS provides higher [CH_4_] values than MIR DCS at high concentrations.

**Table 1.**
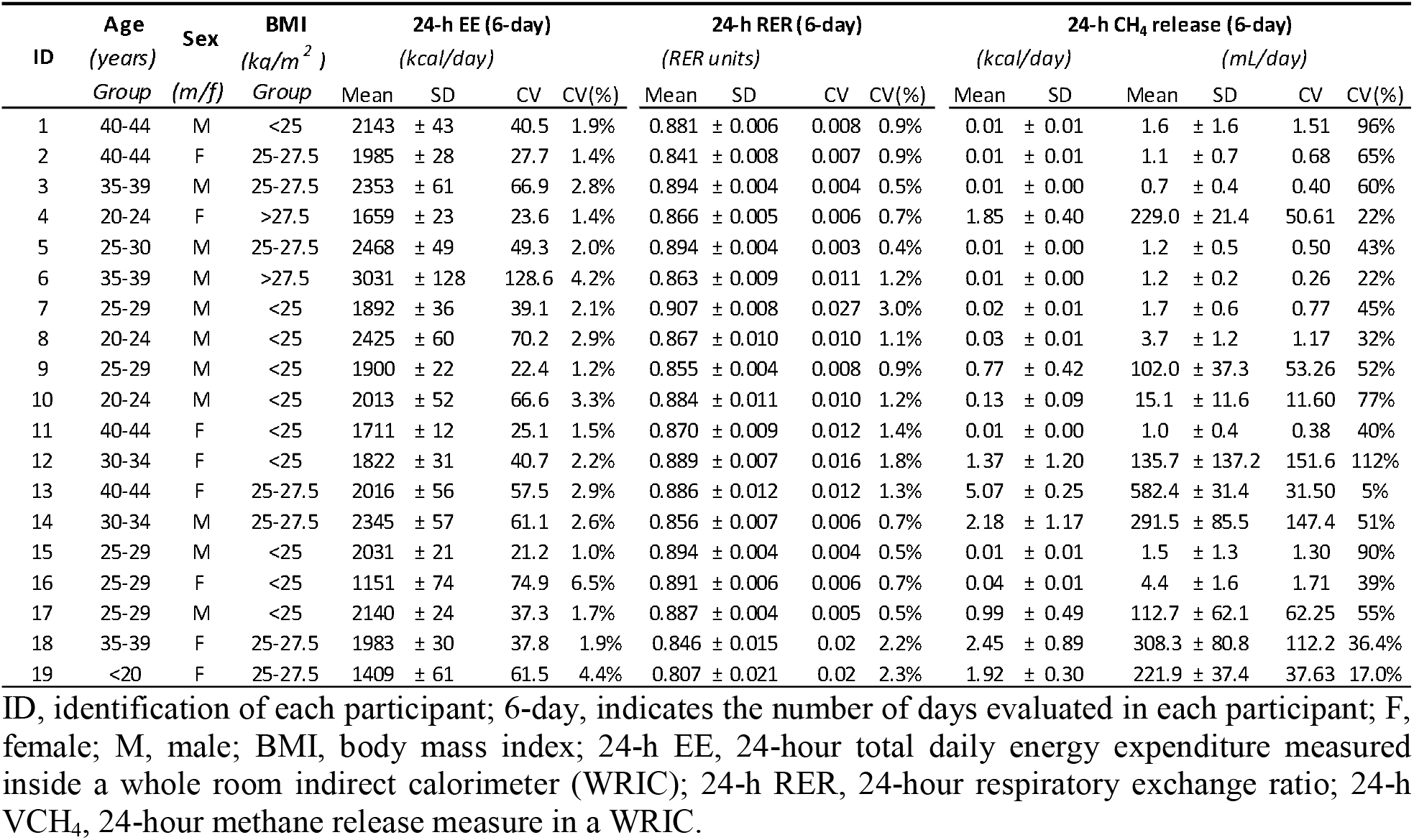
Demographic and metabolic characteristics of study participants.

**Figure 4.**
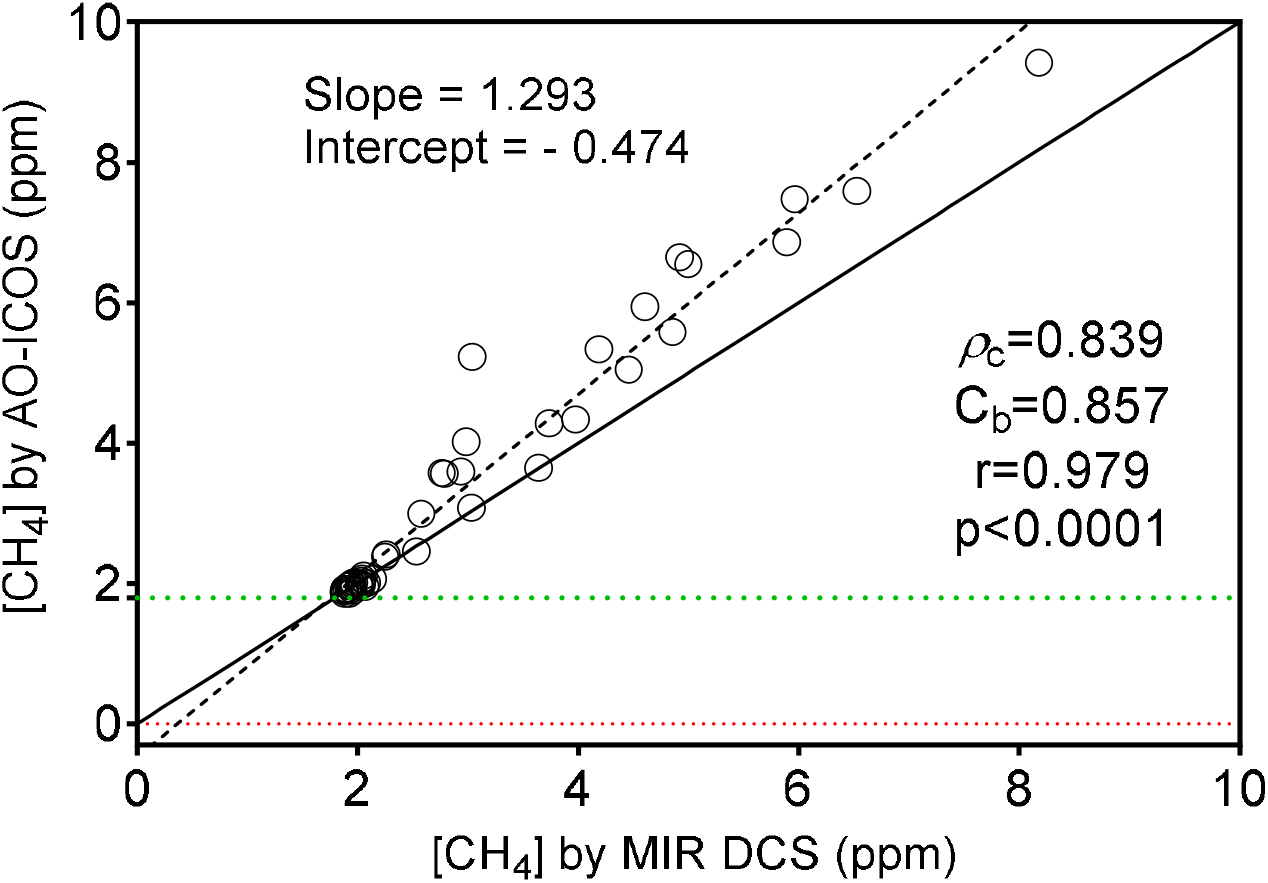
Concordance Correlation Coefficient diagram between methane concentration ([CH_4_]) measured by broadband mid-infrared laser spectroscopy (MIR DCS) and off-axis integrated-cavity output spectroscopy (OA-ICOS). Solid line is the identity line (45 degrees); dashed line represents regression between [CH_4_] measured by OA-ICOS and MIR DCS. ρ_c_ = concordance coefficient of correlation; Cb= bias correction factor; r, precision. Green dotted line marks [CH_4_] atmospheric (1.8-1.9 ppm). Red line marks the lowest limit of detection for each methodology (AO-ICOS = 0.003 ppm and MIR DCS = 0.0001 ppm).

**Figure 5.**
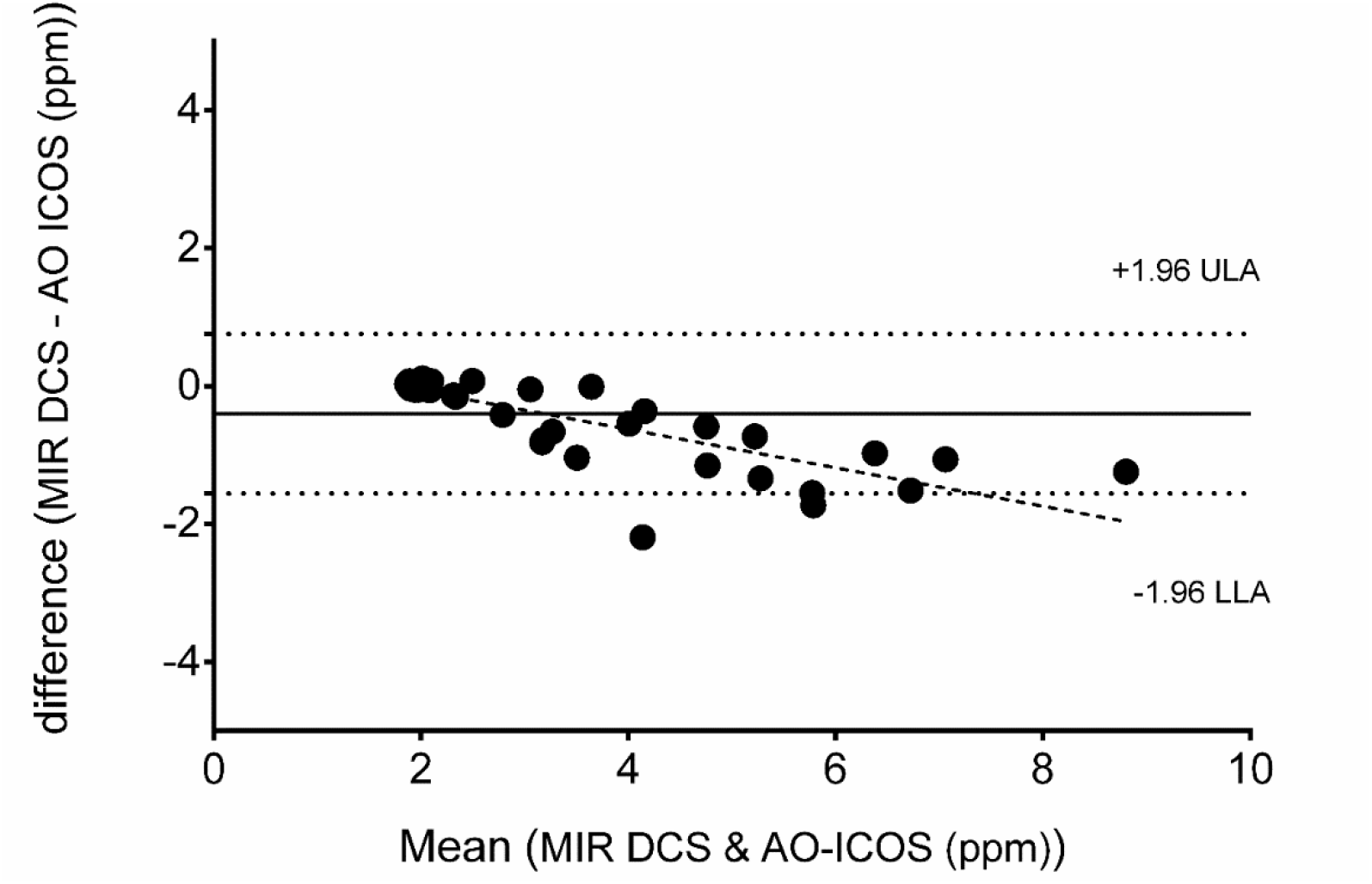
Bland and Altman plot for the agreement analysis between CH_4_ concentration ([CH_4_]) measured by broadband mid-infrared laser spectroscopy (MIR DCS) and off-axis integrated-cavity output spectroscopy (OA-ICOS). Solid line represents mean difference between methodologies (−0.4002 ppm); dotted lines are upper (ULA) and lower (LLA) limit of agreement (ULA: 0.7576 and LLA: –1.558, respectively); dashed line is the mean simple regression line between differences and mean values as indicator of proportional bias.

### Data Processing and Analysis to Differentiate Breath from Colonic Released VCH_4_

Stool mass produced per patient did not correlate with VCH_4_ (mL/min); although, we could observe significant peaks of VCH_4_ during/after each period of stool collection inside the WRIC (Figure 6a). Therefore, the absolute mass of stool could not be considered as a predictor of 24-hour VCH_4_, but an instantaneous indicator of CH_4_ production, at least in the sample of participants included in this study.

**Figure 6.**
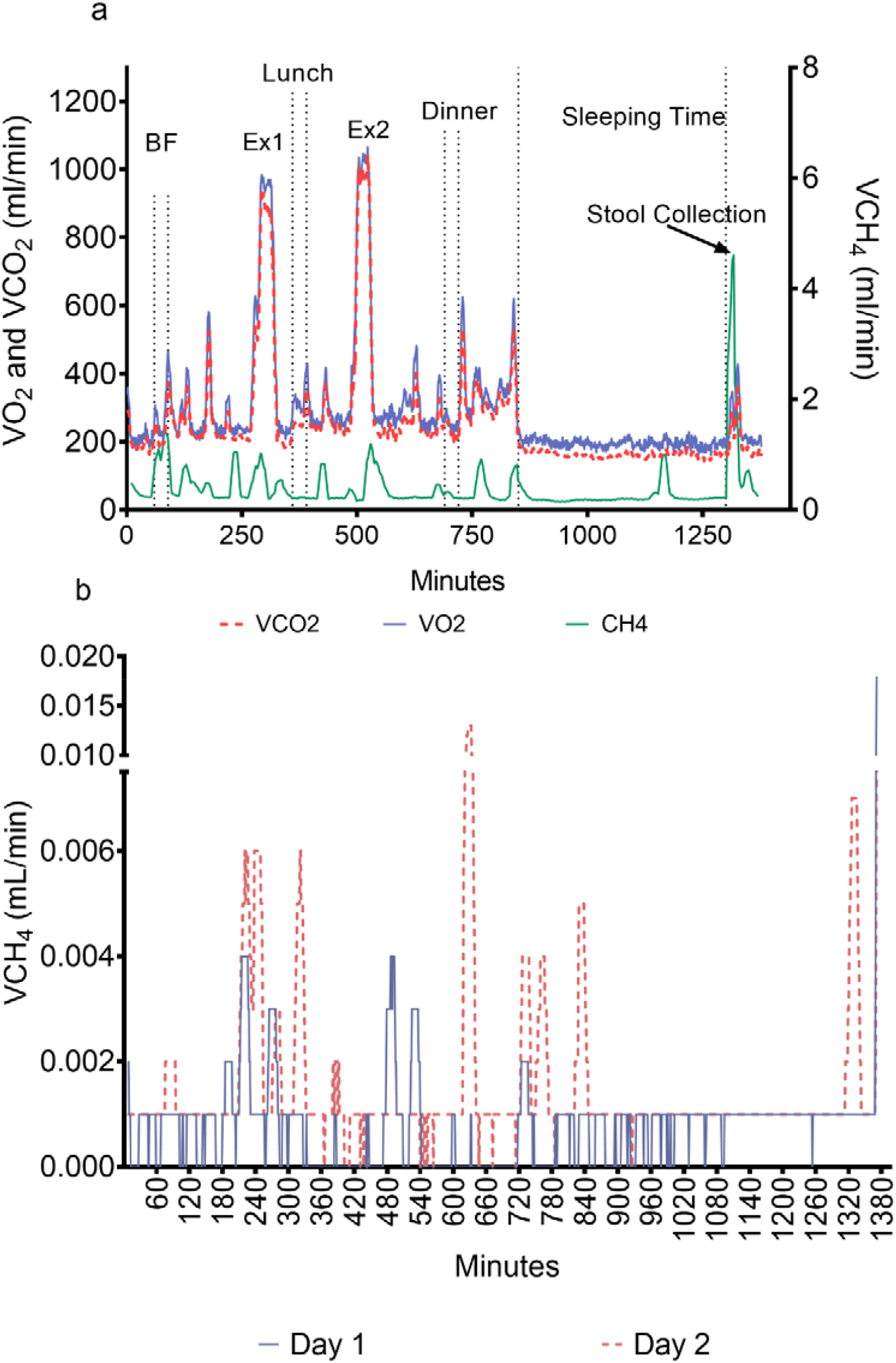
Profiles of 23-h VCH_4_ release in humans living inside a whole room indirect calorimeter (WRIC). Figure 6-a shows simultaneous measurement of VCH_4_, VO_2_ and VCO_2_ from a participant during a single day. Vertical dotted lines split scheduled activities: BF, breakfast time; EX, exercise. Grey solid line is VO_2_ profile; black dashed line is VCO2 profile; black solid line is VCH_4_ profile. Figure 6-b shows VCH4 profiles of the same participant during two consecutive days (solid blue and red dashed lines represent day 1 and 2, respectively).

Figure 6b provides an example of the day-to-day VCH_4_ variability. The between-day reproducibility of 24-hour methane release in humans was highly variable (6-day overall CV=46.4 mL/min (43.7%)) ranging from CV= 5% to 112% (table 1). The overall between-week CV=146.6 ml/day (95.7%). Also, the range of individual variability differs between in days without stool production (CV= 7% to 66%) and days with stool collections (CV= 7% to 127%). The high within and between subject variability of CH4 release contrasts with the low variability of 24-h EE and RER (Table 1).

Our data indicate that 55.8±20.6% of 24-hour CH_4_ was released by breath (48.1±73.52mL/day) when considering all days with and without stool collection inside WRIC. In the days with stool collection outside and inside of WRIC, stool collection only outside of WRIC and without any collection, estimations of 24-hour CH_4_ released by breath were similar: 55.1±20.3% (47.5±73.0 mL/day), 54.8±20.4% (48.2±73.3 mL/day) and 53.0±20.5% (42.2±71.3 mL/day), respectively.

## Discussion

After a series of experiments and validation studies we were able to implement a system to measure VCH_4_ continuously in parallel with a WRIC. This system will be utilized in future studies to calculate EE with more precision by including the VCH_4_ in the stoichiometric equations (3), providing information about acute responses of the fermentation process inside the colon and serving as a template to assess other VOC.

### Capacity of the Technology to Measure CH_4_ Inside WRIC

We used an OA-ICOS analyzer to measure metabolic [CH_4_]. This system was easy to use and install, low power and rugged. It has many advantages over conventional optical techniques (including first generation cavity ringdown spectroscopy) such as being simpler to build, more rugged and alignment insensitive, having a much shorter measurement time, and providing measurements over a much wider dynamic range. However, our studies revealed important aspects to be considered during the installation of this type of system. For example, the environmental context of the facility is a key factor when installing the pipeline to collect the medical air that will feed the WRIC with the inflow air. Environmental fluctuations of CH_4_ dramatically affect CH_4_ production calculations. Therefore, an initial analysis of the weekly [CH_4_] variability around the facility is highly recommended as the first step before the installation of the gas collection pipe.

The second step before coupling the CH_4_ analyzer with the WRIC system, was to cross-validate [CH_4_] values of the OA-ICOS with an independent system (MIR-DCS). Our data confirmed that both analyzers measured similar values and no CH_4_ was lost between the WRIC and the analyzer. After this initial experiment of cross-validation, we integrated the measurements of the OA-ICOS with the WRIC outputs (VO_2_ and VCO_2_), which required calculating the volumes with MFC and aligning time of the measurements of three gases. Specific code was written in Excel (Supplemental Material), which was able to provide VCH_4_ data with a 2-minute resolution. To validate this new setup, we performed infusion studies with a blender system in the two large WRICs in our facility. The short-circuit infusion validated the detection of VCH_4_ signals at progressively increasing volume. The static infusions revealed that the system was able to provide valid and reliable VCH_4_ assessments at variable volume outputs. The reliability between days and weeks was high for the same chamber; however, the studies between chamber showed a larger difference.

### Cross-Validation and Measurements of Human Samples

Seminal studies by the Marinos Elia’s group in the United Kingdom constituted the first attempt to capture 24-hour VCH_4_ production (7, 18); however, these studies collected samples of humans every two hours and only in sedentary conditions. We are the first group reporting continuous data at high resolution (2-minute) inside a WRIC without movement restrictions. Single-time point [CH_4_] measurements during several 24-hour periods correlated well with another independent technology (MIR DCS), which confers external cross-validation to our assessments. Nevertheless, we found a proportional bias where higher VCH_4_ produced a larger the difference between OA-ICOS and MIR DCS (lower [VCH_4_] were measured with the MIR DCS). This phenomenon could have been due to our gas sampling method to collect gas from the pipeline of WRIC. We cannot rule out the possibility of small leaks from the gas sampling bags, which has been previously reported in other studies analyzing VOCs in human breath with a similar collection method (19).

The reliability studies in humans of the 24-hour VCH_4_ showed a high intra-day and between-day variability, which was not correlated with the volume of feces; this is likely due to biological variation in the population size and activity of methanogenic archaea per stool mass (12). Significant methane peaks were identified and associated with the collection of stools. This constituted an additional construct to validate response of our system. Nonetheless, the peaks associated with the moment of stool collection were not the only ones. There were significant peaks above the steady state, which may be associated with flatulence and constitutes the second source of intestinal methane.

### Differentiation Between CH4 Released Through Breath and Intestine

Our human data confirm results from single breath studies and partial 24-hour production that reported large VCH_4_ variability between participants (11). Therefore, it has been described that diet, disease and specific microbiome composition must be determinants of this variability (10, 12, 18). However, it is not clear if this variability is due to increases in breath or colonic CH_4_ released and/or the proportion that each of them represents to the 24-hour VCH_4_.

We designed a postprocessing analytical method to differentiate colonic and breath CH_4_ release that will help to interpret physiological adaptations in future studies. Our starting paradigm relied on the fact that CH_4_ is produced by archaea in the colon and is not significantly produced or consumed by the human body after entering in the portal circulation from the colon. Also, there are several behavioral events that promote the release of intestinal gas on daily basis (walking, abdominal bending, eating/digestion or type of diet), which may be inhibited during sleep. Bearing this in mind, it seems reasonable that any volume of CH_4_ significantly higher than the normal variation (CV of technology + CV while resting/sleeping) should be considered as colonic release. Our data indicated that intestinal CH_4_ release was below 50%, which is lower than the traditional values of 80% reported in the literature (10). Nonetheless, our method must be validated in a future study.

There are sources of error that could have affected our human studies. While certain states of disease and medications/supplements can affect CH_4_ production (10-12, 18), we were not able to address this in this study since all participants needed to be healthy and not taking any medication that could affect energy metabolism. Some of the participants could have modified their movement and normal toilet activities due to the constrained and unusual environment inside the WRIC. Therefore, some did not go to the toilet in the WRIC but during the daily one-hour outside the WRIC. Since we observed significant VCH_4_ peaks during stool collection, this could have underestimated the total CH_4_ production in some cases. We were not able to create an equation to estimate CH_4_ production outside of the WRIC utilizing our current dataset and variables.

## Conclusion

Our results suggest that the use of OA-ICOS device integrated in a WRIC setup provides valid and reproducible assessments of VCH_4_. We also suggest using the same WRIC in longitudinal studies to minimize the error associated with the technique. Postprocessing data to differentiate intestinal from breath CH_4_ release along with its physiological implications remain an open area of study and a challenge for future experiments. Also, high CH_4_ variability between days in humans requires further study as it is a biological phenomenon that is not likely due to limitations of highly controlled studies that do not mirror a real-world environment. The measurements of small volatiles and inorganic molecules inside a WRIC in highly controlled conditions is a critical first step to establish testable hypotheses for free-living conditions.

Therefore, this new system will facilitate the study of new hypotheses such as the impact of different types of diets, supplements or any microbiome-targeting interventions on CH_4_ release and its association with energy metabolism, intestinal physiology, disease states and microbiome function.

## Data Availability

All data produced in the present study are available upon reasonable request to the authors

## Acknowledgments

We thank the volunteers who provide the breath and biological samples for the human validation studies. We also thank the staff of all institutions involved in the assessments and studies. Research reported in this publication was supported by the National Institute of Diabetes and Digestive and Kidney Diseases of the National Institutes of Health under Award Number NCT02939703. The content is solely the responsibility of the authors and does not necessarily represent the official views of the National Institutes of Health

