## Supplemental figures for "Measurement of 24-hour Continuous Human CH_4_ Release in a Whole Room Indirect Calorimeter"

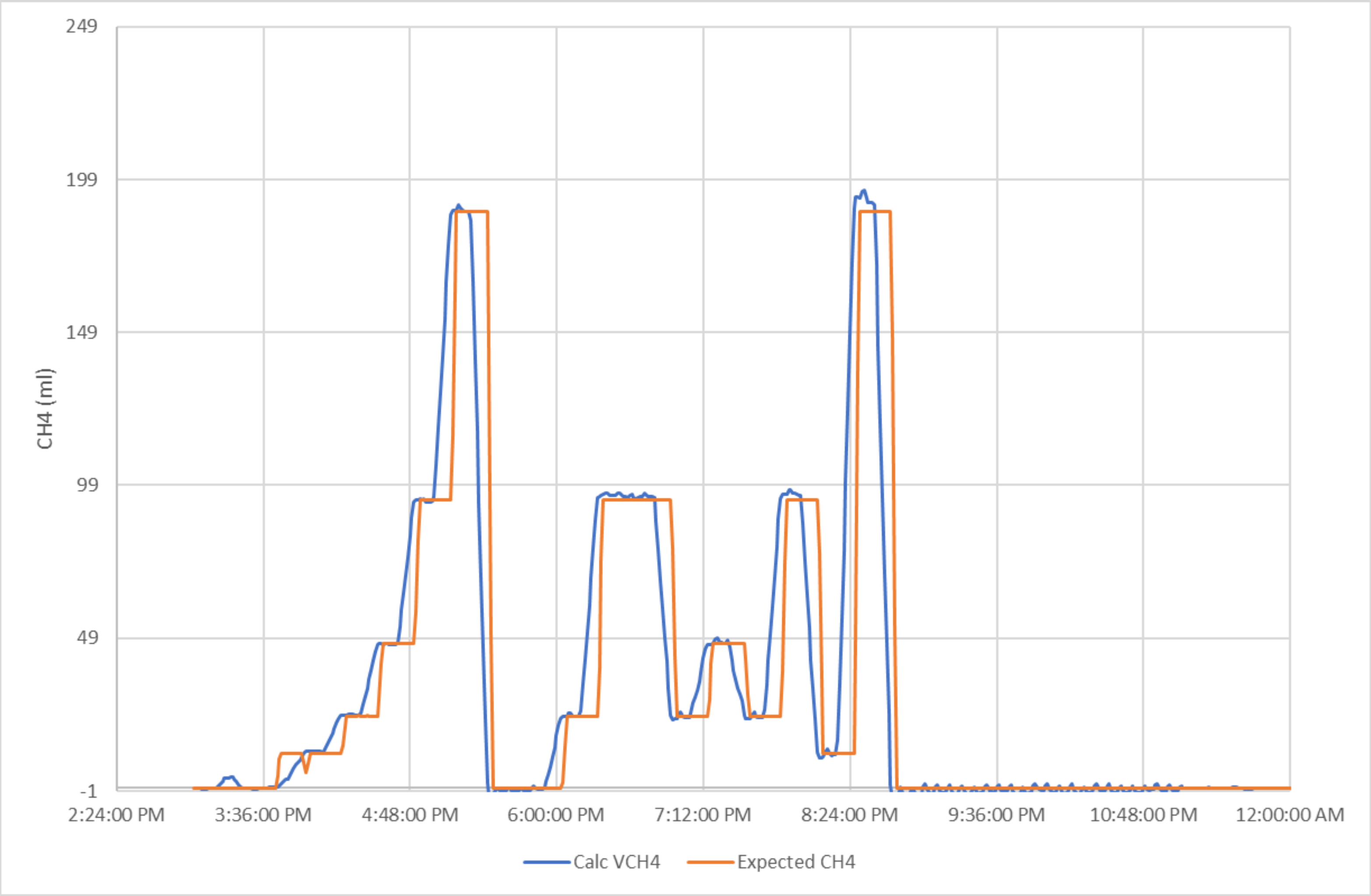


**Supplemental Figure 1**. Short step CH4 infusion. First section of the graph shows time progression infusions (10 minutes/per step). Second section of the graph shows time random variable infusions (10 minutes per step).

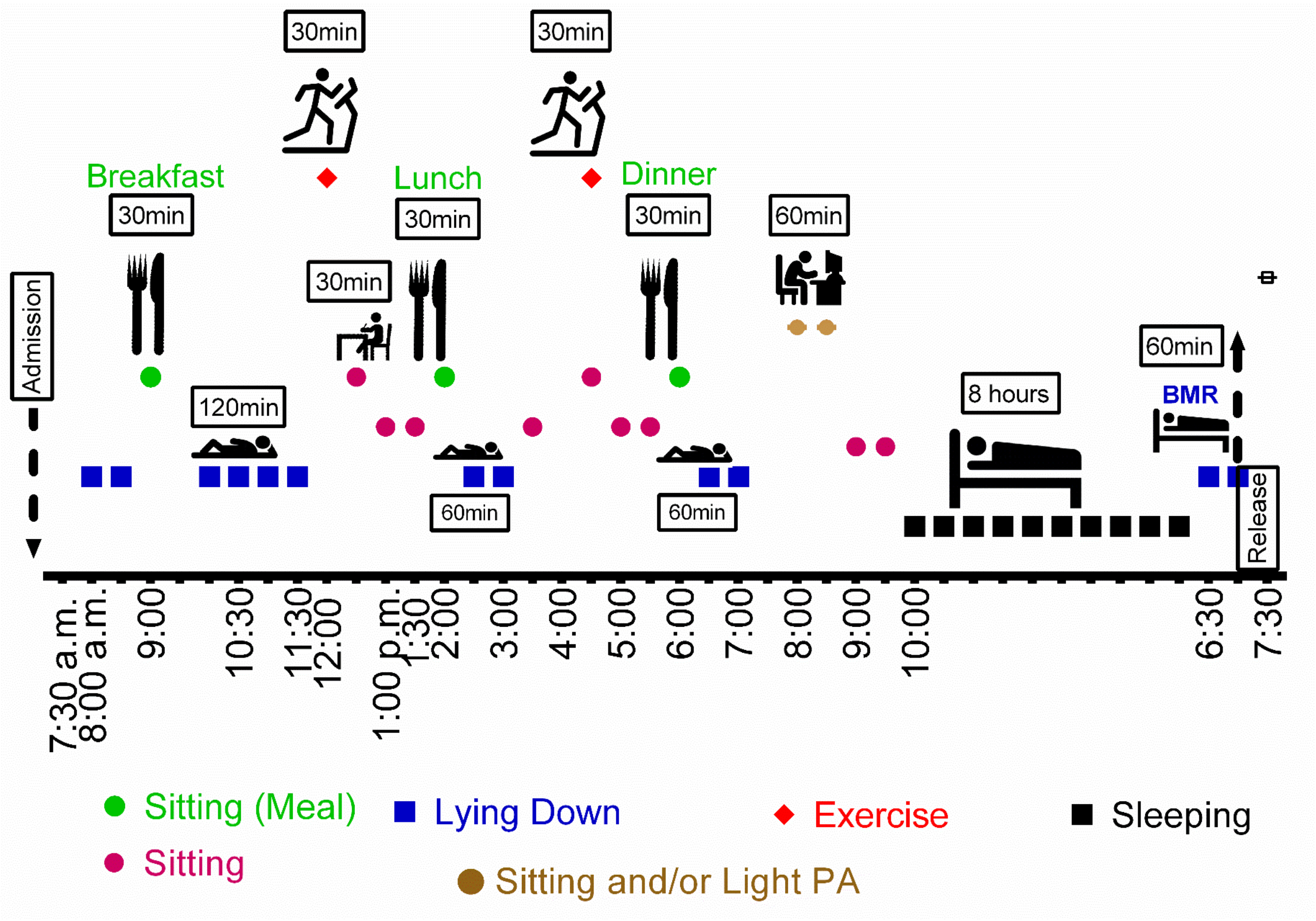


**Supplemental Figure 2**. Schedule of events inside the whole room indirect calorimeter (WRIC).

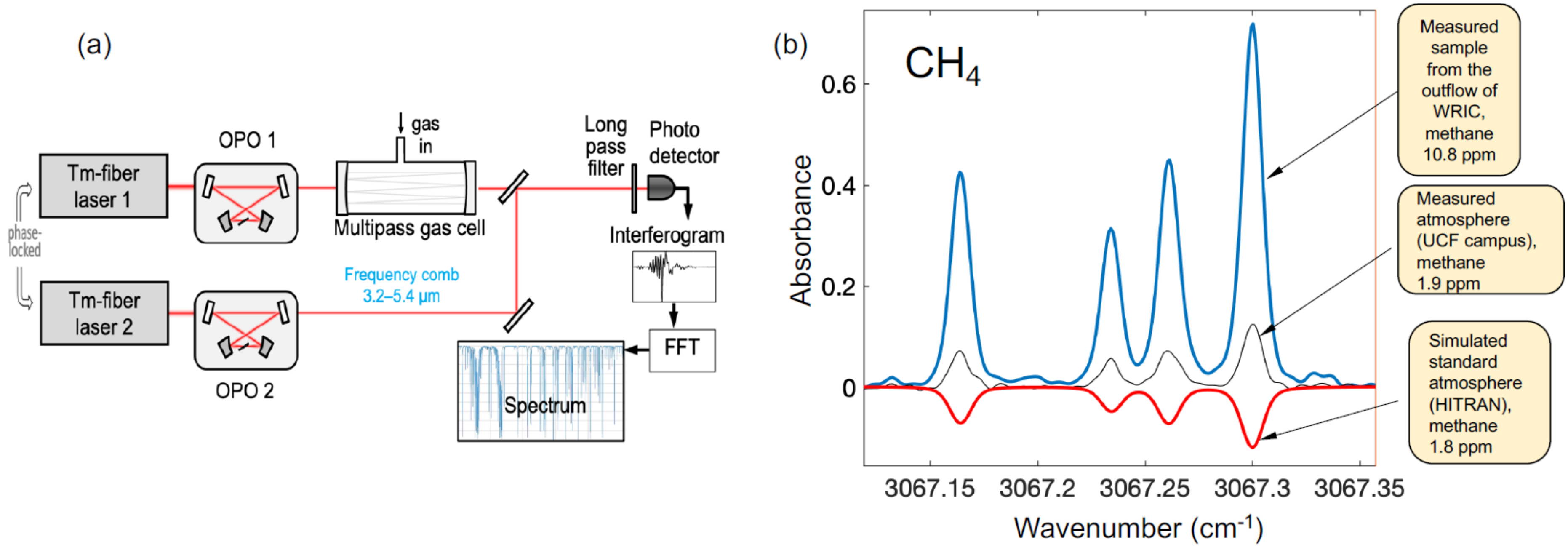


**Supplemental Figure 3.** (a) Schematic of the MIR DCS setup. A pair of phase-locked Tm-fiber lasers pump a pair of subharmonic OPOs. The two beams are combined, passed through a multipass gas cell with an equivalent path length of 76 m, and detected with a fast infrared detector. The detector signal is digitized, and fast Fourier transformed to retrieve the optical spectrum. (b) Measured and simulated absorption spectra of $CH_4$. Measured are examples of sample from whole room indirect calorimetry (WRIC) at the translational research institute (TRI) and University of Central Florida (UCF) A simulated spectrum (HITRAN) is inverted for clarity.
